# Performing moderate to severe activity is safe and tolerable for healthy youth while wearing a cloth facemask

**DOI:** 10.1101/2022.03.11.22272270

**Authors:** Felipe Miguel Marticorena, Gabriel Barreto, Natália Mendes Guardieiro, Gabriel P. Esteves, Tamires Nunes Oliveira, Luana Farias de Oliveira, Ana Lucia de Sá Pinto, Luiz Riani, Danilo Marcelo Leite do Prado, Bryan Saunders, Bruno Gualano

**Affiliations:** Applied Physiology and Nutrition Research Group, School of Physical Education and Sport; Rheumatology Division; Faculty of Medicine FMUSP, University of São Paulo, São Paulo, SP; Clinical Hospital, Faculty of Medicine FMUSP, University of São Paulo; Laboratory of Assessment and Conditioning in Rheumatology, Rheumatology Division, Universidade de São Paulo, São Paulo, Brazil; Institute of Orthopaedics and Traumatology, Faculty of Medicine FMUSP, University of São Paulo, Brazil; Food Research Center, University of São Paulo, São Paulo, Brazil

**Keywords:** physical activity, mask, COVID-19 pandemic, oxygen saturation, lactate

## Abstract

**Objective:** To investigate if wearing a cloth facemask could affect physiological and perceptual responses to exercise at distinct exercise intensities in healthy young individuals.

**Methods:** In a crossover design, 9 participants (sex, female/male: 6/3; age: 13±1 years; BMI: 18.4±2.1 kg/m^2^; sexual maturity rating, I/II/III/IV: 0/3/4/2; VO_2peak_: 44.5±5.5 mL/kg/min) underwent a progressive square-wave test at four intensities: (1) at 80% of the ventilatory anaerobic threshold (VAT), (2) at VAT, and (3) at 40% between the VAT and V□O_2peak_ wearing a triple-layered cloth facemask or not. These stages represented moderate, heavy, and very heavy domains and corresponded to 46±8%, 57±10% and 87±8% of V□O_2peak_. Participants then completed a final stage (severe) to exhaustion at a running speed equivalent to the maximum achieved during the cardio-respiratory exercise test (Peak). Physiological, metabolic, and perceptual measures were analysed.

**Results:** Mask did not affect spirometry (forced vital capacity [FVC], peak expiratory flow [PEF1], forced expiratory volume [FEV]; all p > 0.27; Figure 1), respiratory (inspiratory capacity [IC], end-expiratory volume to functional vital capacity ratio [EELV/FVC], EELV, respiratory frequency [Rf], tidal volume [VT], Rf/VT, end-tidal carbo dioxide pressure [PetCO_2_], ventilatory equivalent and carbon dioxide ratio [VE/VCO_2_]; all p > 0.196), hemodynamic (heart rate [HR], systolic and diastolic blood pressure [SBP; DBP]; all p > 0.41), rated perceived exertion (RPE; p = 0.04) or metabolic measures (lactate; p = 0.78 Figure 2) at rest or at any exercise intensity. In both conditions, the same number of children (4 out of 9) were unable to finish the Peak stage, whereas one child did not complete exercise at the heavy domain while wearing no mask.

**Conclusions:** This study shows that performing moderate to severe activity is safe and tolerable for healthy youth while wearing a cloth facemask. ClinicalTrials.gov: NCT04887714

## Introduction

Mask mandates became one of the most important measures to prevent infections during the COVID-19 pandemic (1-3). Wearing a facemask can be particularly relevant at schools, indoor gyms, and fitness centres where outbreaks have been identified (4, 5). However, concerns have been raised regarding the safety and tolerability of wearing masks while exercising (6).

Among adults, facemasks (cloth, surgical) and respirators (FFP2/N95) have been shown to reduce the ability to breathe comfortably during exercise, which has been confirmed by some (7, 8), but not all (9) studies. Recently, we demonstrated that a cloth facemask did not cause major perturbations in respiratory or cardiovascular responses during moderate to heavy exercise performed by non-trained men and women, although time-to-exhaustion at maximal intensity was decreased while wearing a mask.

Cumulative evidence now suggests that performing physical activity is safe and tolerable for adults while wearing a facemask (9). However, little is known on the impact of facemasks in the youth. Marked differences exist in the physiological responses to exercise between children and adults as supported by classical studies. For instance, children have lower stroke volume than adults at all levels of exercise intensity, which is partially compensated by a higher heart rate (10). This results in slightly lower cardiac output during exercise, possibly affecting oxygen transport system, particularly at near maximal and maximal intensities when peripheral oxygen extraction reaches a plateau (11, 12). In line with the lower cardiac output and stroke volume is a lower exercise systolic and diastolic blood pressure in exercising children (13, 14). In addition, compared to young adults, children respond to exercise with relative tachypnea, shallow breathing, higher respiratory frequency and ventilatory equivalent (which reflects lower efficiency) (15), and increased perceived effort, at least in more prolonged exercise (≥ 15 min) (16, 17).

Hypothetically, the less efficient cardiorespiratory responses and the higher perceived effort in exercising children could be aggravated by wearing a mask, particularly at higher intensities. This study aimed to test whether a facemask could affect physiological and perceptual responses to exercise healthy youth at different intensity domains, determined by a progressive square-wave test (PSWT).

## Methods

### Ethics statement

The protocol was approved by the institutional ethics committee. Written informed consent/assent was obtained from the parents/children prior to participation.

### Study design and setting

This was a crossover study (clinicaltrials.gov: NCT04887714) performed at an intrahospital exercise physiology laboratory. Data collection took place between November 2021 and January 2022.

### Participants

Healthy children of both sexes were eligible for this study. Exclusion criteria included any cardiac, pulmonary, and rheumatologic diseases, musculoskeletal limitations, or a BMI >30 kg·m^2^. A total of 10 children participated in the study; 1 child dropped out for personal reasons. Nine participants completed all main sessions and were analysed. Participant characteristics are presented in Table 1. All children were physically active, according to the Physical Activity and Sedentary Lifestyle Assessment Questionnaire for Children and Adolescents (18).

**Table 1.**
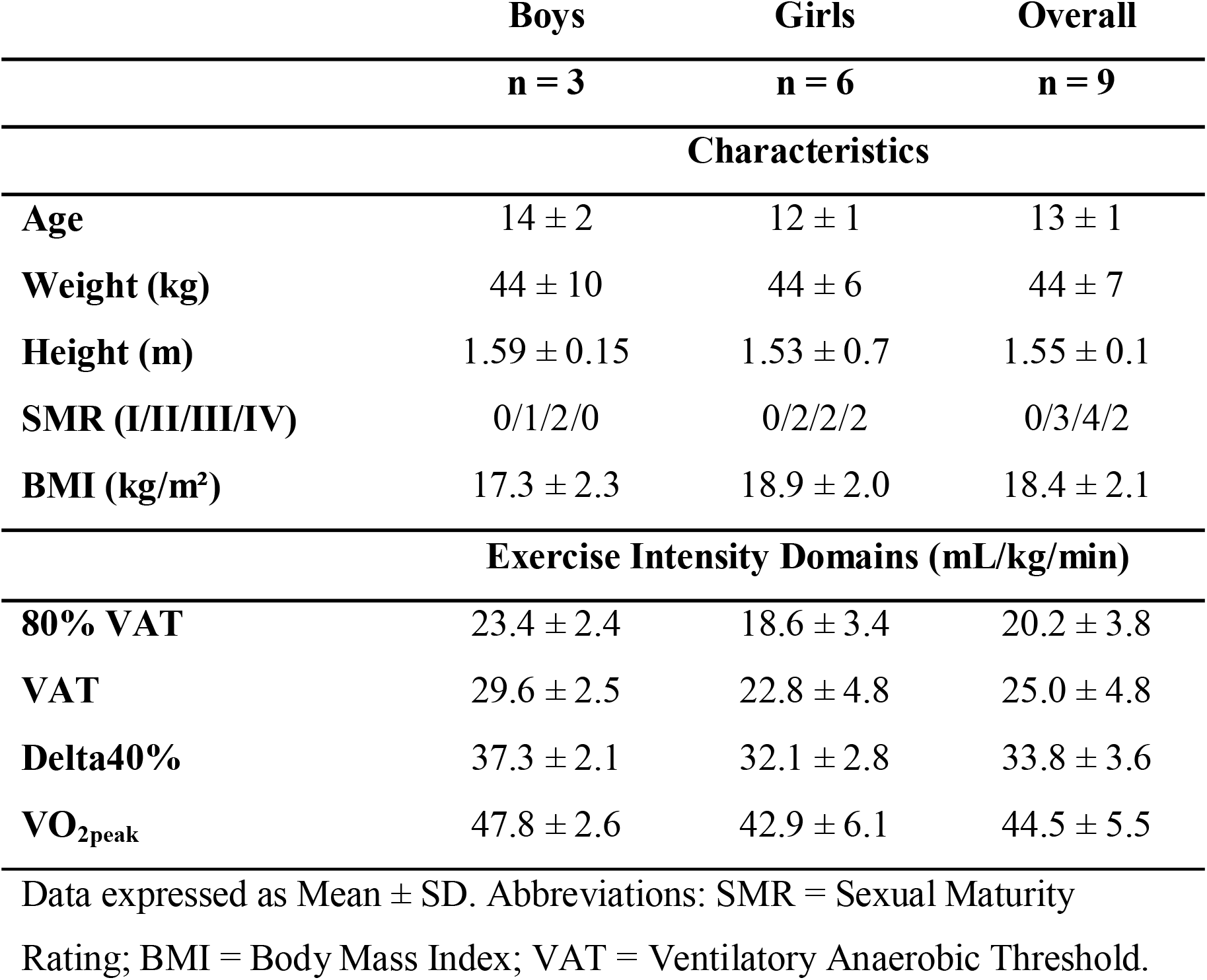
Participant characteristics.

### Experimental design

Participants attended the laboratory on four separate occasions, separated by a minimum of 48 h, at the same time of day to account for circadian variation (19). The first visit consisted of an incremental cardiopulmonary running test to exhaustion to determine peak oxygen uptake (V□O_2peak_) and ventilatory thresholds. The second visit was for a familiarization session to the PSWT without wearing a mask. The remaining two main visits consisted of the PSWT performed with or without the use of a triple-layered antiviral cloth facemask (Fashion Masks, São Paulo, Brazil). This facemask was chosen because it is widely accessible, recommended to the general public by the CDC and appropriate for exercise (https://www.cdc.gov/coronavirus/2019-ncov/prevent-getting-sick/cloth-face-cover-guidance.html). The outer layer was waterproof polyester fabric, the middle layer a polypropylene filter, and the inner layer was absorbable cotton. The size was “one size fits all” and was thus identical for all participants. Participants were required to keep the mask in place over the nose, mouth, and chin during the entire session. The breath-by-breath facemask was placed over the cloth facemask, and participant were required to exhale as forcefully as possible while blocking the inlet/outlet hole, allowing the researchers to adjust the mask to ensure minimal air escaping. The order of sessions was determined by an individual not involved in data collection. Blocks of two individuals were allocated to the two possible orders (Mask– No Mask; No Mask–Mask) using a random number generator (https://www.randomizer.org/) to ensure the study was counterbalanced. Since the investigators could see that the participants were wearing a cloth facemask or not, the session order was provided directly to the research team. All participants were habituated to wear a mask during their daily routines due to mandates, but not specifically during exercise. Participants were requested to refrain from strenuous exercise, caffeine, and replicated their diet, in the 24 h prior to each visit.

### Cardiorespiratory Exercise Test

Immediately prior to the cardiorespiratory exercise test, participants performed a pulmonary function test according to previous recommendations (20). The cardiorespiratory exercise test was performed on a motorized treadmill (Centurion 300, Micromed, Brazil). The test started at 4 km·h^-1^ and increased speed (1 km·h^-1^·min^-1^) up to a maximum velocity of 14 km·h^-1^. For those participants who reached these maximal speeds, there was a subsequent increase in inclination (2%·min^-1^) until exhaustion. Ventilatory and gas exchange measurements were recorded continuously throughout the test using a breath-by-breath system (MetaLyzer 3B, Cortex, Germany), as was heart rate (HR; ergo PC elite, Micromed, Brazil). Maximal effort was determined according to published criteria and individual V□O_2peak_ was determined as the V□O_2_ averaged over the final 30 s.

### PSWT

To determine exercise workload for the PSWT, data from the cardiorespiratory exercise test were used. All exercise intensity domains were determined by the same respiratory physiologist with experience in the area. The protocol was performed on a motorized treadmill (Centurion, model 200, Micromed, Brazil) and consisted of three 5-min stages at workloads equivalent to (1) 80%VAT, (2) VAT, and (3) 40% of the difference between VAT and V□O_2peak_ (40%Δ). These stages represented moderate, heavy and very heavy domains (21) and corresponded to 46±8%, 57±10% and 87±8% of V□O_2peak_. Participants then completed a final stage (severe domain) to exhaustion at a running speed equivalent to the maximum achieved during the cardiorespiratory exercise test (Peak). Ventilatory and gas exchange measurements were recorded continuously throughout using a breath-by-breath system (MetaLyzer 3B, Cortex, Germany), with the spirometer mask placed over the cloth facemask.

To determine the effect of the mask on pattern of change in operating lung volume, we evaluated end-expiratory volume to functional vital capacity ratio (EELV/FVC). Inspiratory capacity was determined at rest and at the end of each exercise stage during the PSWT. Ventilatory constraint was evaluated as the difference between inspiratory capacity at rest and at each exercise workload (22). Ventilatory efficiency was determined using the ventilatory equivalent for carbon dioxide (V□_E_/V□CO_2_) and end-tidal carbo dioxide pressure (PetCO_2_) during each stage. Breathing pattern was evaluated during each stage using the breathing frequency to tidal volume ratio (Rf/VT) ratio (23).

Rated perceived exertion (RPE) was assessed at the end of each stage with participants pointing to a chart using the 6-to 20-point Borg scale (24). Heart rate was monitored continuously throughout (ergo PC elite, Micromed, Brazil). A fingertip blood sample (20 μL) was collected at baseline, at the end of each stage and 4-min post-exhaustion for the subsequent analysis of lactate. Blood was homogenized in the same volume of 2% NaF, centrifuged at 2000 g for 5 min before plasma was removed and stored at -20°C until analysis. Plasma lactate was determined spectrophotometrically using an enzymatic-colorimetric method (Katal, Interteck, Brazil).

### Statistical analyses

Repeated measures Mixed Model ANOVAs were performed with condition (Mask, No-Mask), and exercise intensity (Baseline [except RPE], 80%VAT, VAT, RCP, Peak) as fixed factors and individuals as random factors. The spirometry variables were not repeated measures and, therefore, time was not included as a fixed factor for these analyses. When a significant main effect or interaction was detected (accepted at p≤0.05), post-hoc pairwise comparisons were performed with Tukey’s adjustment. Lactate data were log10 transformed before mixed model analysis, turning the model into an exponential data mixed model, and back transformed through exponentiation for the final reporting of data. Estimated mean changes were extracted from the mixed model. Whenever outlying data points were believed to be improbable (e.g., a value of 50 mmHg for systolic blood pressure), they were considered measurement or transcription error and were excluded. All analyses were performed with RStudio software (Rstudio 1.4.11003, PBC, Boston, MA) using the “lmer” function from the lmerTest package, and the “emmeans” function from the emmeans package. Standard errors were transformed into 95% confidence intervals (CI). All values are expressed as estimated differences and 95%CIs, and data in figures are represented as mean ± 1 standard deviation.

## Results

Mask did not affect spirometry (FVC, PEF1, FEV; all p > 0.27; Figure 1), respiratory (IC, EELV, EELV/FVC, Rf, VT, Rf/VT, PetCO_2_, VE/VCO_2_; all p ≥ 0.196; Table 2), hemodynamic (HR, SBP, DBP; all p ≥ 0.41; Table 2), or metabolic measures (lactate; p = 0.78, Figure 2) at rest or any exercise intensities. An effect of Mask was detected for RPE (condition*intensity interaction: F = 2.93, p = 0.04). Nonetheless, post-hoc analyses did not detect any significant differences between conditions at any exercise intensity (Figure 2). In both conditions, the same number of children (4 out of 9) were unable to finish the Peak stage, whereas one child did not complete the Delta40% domain in the No-Mask condition.

**Figure 1.**
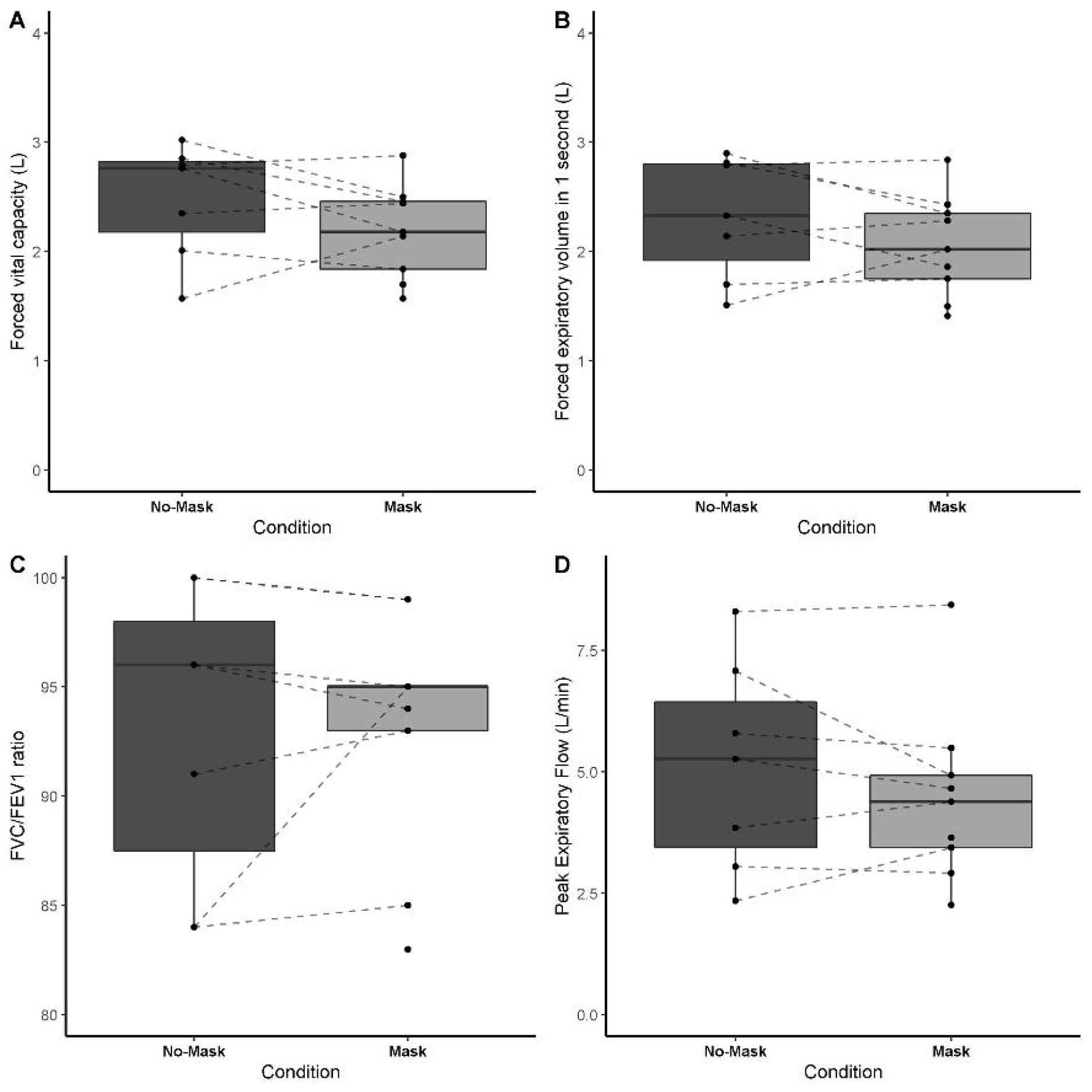
Spirometry data at rest with or without wearing a facemask. All data are expressed as mean and standard deviations. No significant differences between conditions were found.

**Figure 2.**
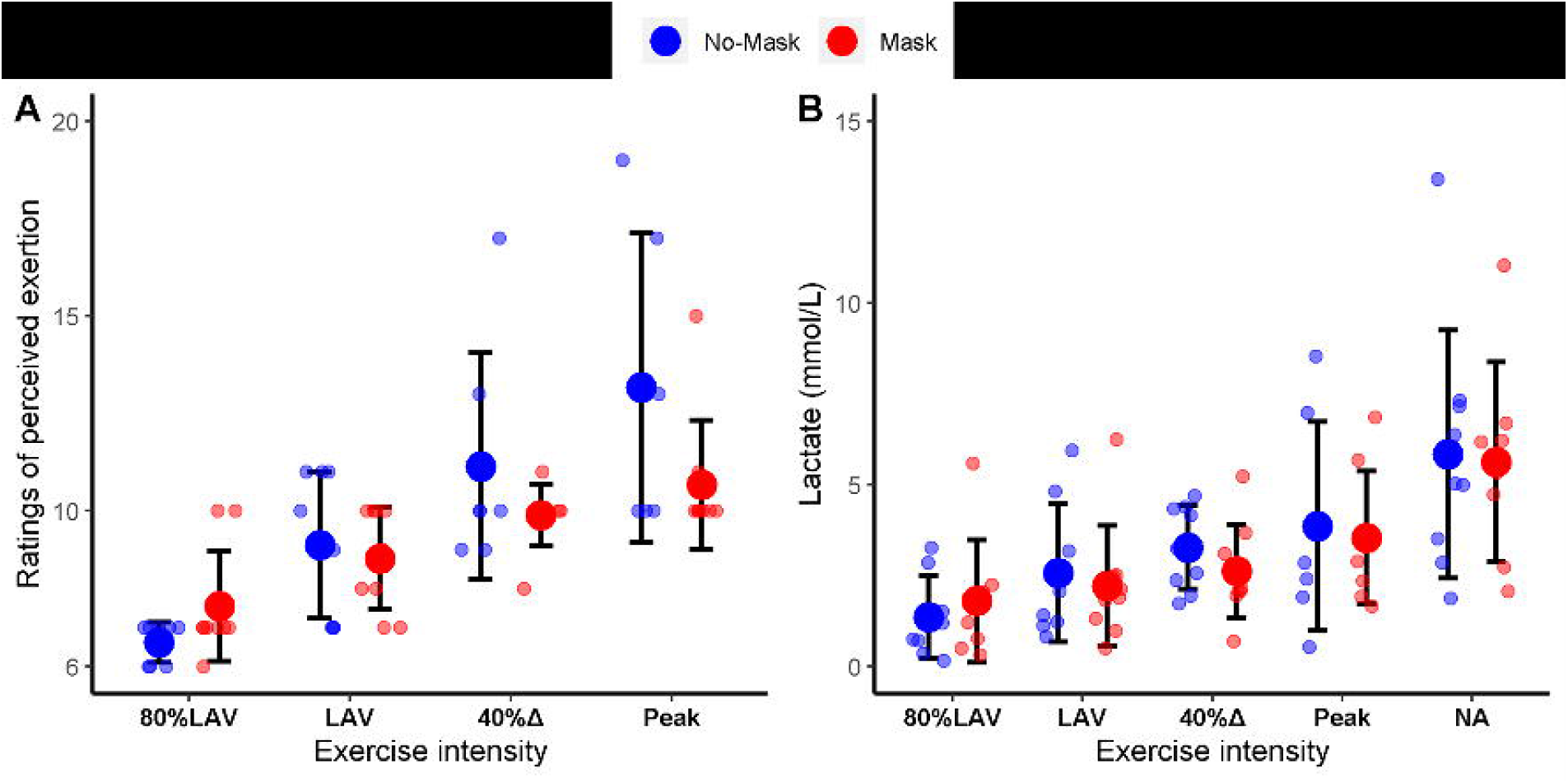
Ratings of perceived exertion (Panel A) and blood lactate concentration (Panel B) during a progressive square-wave test with or without wearing a facemask. Data expressed as means and standard deviations and individual data.

**Table 2.** Data from progressive square-wave test with or without wearing a facemask.

| | Intensity | No-Mask<br>(mean ± SD) | Mask<br>(mean ± SD) | Estimated<br>differences | 95%CI | p-value for<br>interaction |
| --- | --- | --- | --- | --- | --- | --- |
| **Inspiratory**<br>**Capacity** | Rest | 1.9 ± 0.5 | 1.5 ± 0.4 | -0.3 | -0.8;0.1 | 0.585 |
| | 80% VAT | 2.0 ± 0.3 | 1.6 ± 0.4 | -0.3 | -0.8;0.1 | |
| | VAT | 2.1 ± 0.3 | 1.6 ± 0.4 | -0.4 | -0.8;0.1 | |
| | 40%$\Delta$ | 1.9 ± 0.2 | 1.7 ± 0.3 | -0.1 | -0.6;0.3 | |
| | Peak | 2.0 ± 0.4 | 1.7 ± 0.3 | -0.3 | -0.7;0.2 | |
| **End-**<br>**Expiratory**<br>**Lung**<br>**Volume** | Rest | 0.6 ± 0.4 | 0.8 ± 0.4 | 0.2 | -0.4;0.7 | 0.759 |
| | 80% VAT | 0.5 ± 0.4 | 0.7 ± 0.2 | 0.2 | -0.4;0.7 | |
| | VAT | 0.4 ± 0.5 | 0.6 ± 0.2 | 0.3 | -0.3;0.8 | |
| | 40%$\Delta$ | 0.6 ± 0.5 | 0.6 ± 0.2 | -0.0 | -0.6;0.5 | |
| | Peak | 0.5 ± 0.5 | 0.6 ± 0.2 | 0.1 | -0.4;0.7 | |
| **EELV/FVC** | Rest | 0.2 ± 0.2 | 0.4 ± 0.2 | 0.1 | -0.1;0.3 | 0.760 |
| | 80% VAT | 0.2 ± 0.1 | 0.3 ± 0.1 | 0.0 | -0.2;0.3 | |
| | VAT | 0.1 ± 0.2 | 0.3 ± 0.1 | 0.2 | -0.1;0.4 | |
| | 40%$\Delta$ | 0.2 ± 0.2 | 0.3 ± 0.1 | 0.1 | -0.2;0.3 | |
| | Peak | 0.2 ± 0.2 | 0.3 ± 0.1 | 0.1 | -0.1;0.3 | |
| **Respiratory**<br>**Frequency** | Rest | 17 ± 5 | 17 ± 7 | 0.1 | -8.2;8.5 | 0.791 |
| | 80% VAT | 27 ± 6 | 24 ± 7 | -2.3 | -10.7;6.0 | |
| | VAT | 33 ± 7 | 31 ± 8 | -2.7 | -11.0;5.7 | |
| | 40%$\Delta$ | 42 ± 11 | 40 ± 10 | -2.1 | -10.5;6.2 | |
| | Peak | 54 ± 10 | 50 ± 9 | -4.2 | -12.8;4.4 | |
| **BF/TV** | Rest | 39.2 ± 19.7 | 37.2 ± 14.9 | -2.1 | -19.1;15.0 | 0.888 |
| | 80% VAT | 33.3 ± 15.3 | 35.9 ± 22 | 2.6 | -14.5;19.6 | |
| | VAT | 33.8 ± 16.5 | 36 ± 18 | 2.3 | -14.8;19.3 | |
| | 40%$\Delta$ | 39.8 ± 22.2 | 44.2 ± 22.8 | 4.4 | -12.6;21.5 | |
| | Peak | 44.2 ± 17.6 | 48 ± 14.9 | 3.9 | -13.7;21.6 | |
| **PetCO2** | Rest | 31.6 ± 4 | 31.4 ± 4.6 | -0.1 | -2.9;2.6 | 0.544 |
| | 80% VAT | 35.1 ± 4.5 | 35.1 ± 4.3 | 0.0 | -2.7;2.8 | |
| | VAT | 34.9 ± 4.2 | 35 ± 4.2 | 0.1 | -2.6;2.9 | |
| | 40%$\Delta$ | 34 ± 4.2 | 35.2 ± 3.6 | 1.2 | -1.5;4.0 | |
| | Peak | 32.6 ± 4.3 | 33.9 ± 3.8 | 1.3 | -1.5;4.1 | |
| **Tidal Volume** | Rest | 0.5 ± 0.2 | 0.5 ± 0.1 | -0.0 | -0.3;0.2 | 0.196 |
|  | 80% VAT | 0.9 ± 0.2 | 0.8 ± 0.2 | -0.1 | -0.4;0.6 |  |
|  | VAT | 1.1 ± 0.2 | 1 ± 0.3 | -0.1 | -0.4;0.1 |  |
|  | 40%Δ | 1.2 ± 0.3 | 1 ± 0.2 | -0.2 | -0.4;0.1 |  |
|  | Peak | 1.3 ± 0.4 | 1.1 ± 0.2 | -0.2 | -0.5;0.0 |  |
| **VE/VCO2** | Rest | 30.6 ± 3.7 | 29.6 ± 4.5 | -1.1 | -4.8;2.6 | 0.962 |
|  | 80% VAT | 28.6 ± 3.7 | 27.9 ± 4 | -0.8 | -4.4;2.9 |  |
|  | VAT | 29 ± 3.6 | 28.5 ± 3.1 | -0.5 | -4.1;3.2 |  |
|  | 40%Δ | 30 ± 3.5 | 28.9 ± 3.3 | -1.2 | -4.8;2.5 |  |
|  | Peak | 32.2 ± 4.1 | 30.7 ± 3.2 | -1.5 | -5.3;2.3 |  |
| **Diastolic blood pressure** | Rest | 71 ± 10 | 72 ± 10 | 1.4 | -14.9;17.7 | 0.410 |
|  | 80% VAT | 79 ± 8 | 71 ± 7 | -8.0 | -25.5;9.5 |  |
|  | VAT | 81 ± 8 | 75 ± 12 | -5.8 | -23.2;11.6 |  |
|  | 40%Δ | 77 ± 10 | 68 ± 26 | -9.1 | -25.8;7.6 |  |
|  | Peak | 75 ± 9 | 75 ± 11 | -0.4 | -16.7;15.9 |  |
| **Systolic blood pressure** | Rest | 106 ± 8 | 112 ± 12 | 3.9 | -21.1;29.0 | 0.873 |
|  | 80% VAT | 120 ± 15 | 119 ± 13 | -3.1 | -28.1;21.9 |  |
|  | VAT | 131 ± 19 | 131 ± 11 | -1.3 | -26.5;23.9 |  |
|  | 40%Δ | 139 ± 28 | 146 ± 24 | 6.0 | -18.2;30.2 |  |
|  | Peak | 151 ± 29 | 150 ± 22 | -0.5 | -24.7;23.8 |  |
| **Heart Rate** | Rest | 84 ± 18 | 78 ± 17 | -6.2 | -23.6;11.2 | 0.834 |
|  | 80% VAT | 107 ± 20 | 99 ± 19 | -7.7 | -25.1;9.7 |  |
|  | VAT | 130 ± 19 | 124 ± 25 | -5.7 | -23.1;11.7 |  |
|  | 40%Δ | 157 ± 22 | 156 ± 22 | -1.1 | -18.5;16.3 |  |
|  | Peak | 186 ± 11 | 186 ± 11 | -1.4 | -19.4;16.6 |  |

Abbreviations: VAT = ventilatory anaerobic threshold; EELV = evaluated end-expiratory volume; FVC = functional vital capacity; BF = breathing frequency; TV = tidal volume; PetCO<sub>2</sub> = end-tidal carbo dioxide pressure; VE = ventilatory equivalent; V̇CO<sub>2</sub> = carbon dioxide.

## Discussion

This study shows that performing moderate to severe activity while wearing a cloth facemask is safe and tolerable for healthy youth.

The benefits of mask mandates to mitigate COVID-19 cases are unequivocal (1-3). This holds particularly true in schools, indoor gyms, fitness centres and exercise classes with little possibility for ventilation and social distancing. Despite unfounded allegations that wearing a mask during exercise is unsafe and intolerable, growing evidence in adult populations have shown the opposite (9). Herein we extend this notion to healthy young individuals by providing evidence that wearing a cloth facemask during exercise does not cause any meaningful perturbations in respiratory, cardiovascular or perceptual variables in children at all exercise intensities.

These findings imply that, during an unmitigated epidemic, wearing a cloth facemask is a sound mitigation measure that should not hinder physical activity levels of children. The comprehensive assessments performed in the present study provide novel evidence that a cloth facemask does not have any meaningful impact on ventilatory efficiency or hemodynamic (e.g., oxygen saturation; arterial blood pressure) responses in healthy youth, but further studies involving children with pulmonary and cardiovascular comorbidities remain necessary.

The strengths of this study include the novel investigation of youth, the use of a constant-load test that allows determining exercise intensities accurately by normalizing the physiological responses to exercise in relation to the gas exchange or blood acid-base profiles (25), and the broad assessments of physiological and perceptual variables. Limitations include the low sample size, the inclusion of only healthy participants, the assessment of a specific type of facemask (i.e., cloth), and the use of a mask for breath-by-breath measures over the cloth facemask may have contributed to thediscomfort felt by the participants and may also have led to some inaccuracies in measurements due to air escaping, despite all the measures taken to avoid it (see Methods).

In conclusion, wearing a cloth mask had no major impact on cardiovascular, respiratory, and perceptual parameters in healthy youth during moderate to severe exercise.

## Supporting information

CONSORT

## Data Availability

All data produced in the present work are contained in the manuscript.

## Acknowledgments

The authors would like to thank all the participants for taking part in this research. The authors received no specific funding for this work. B.G (2017/13552-2), G.B. (2020/12036-3), G.P.E. (2020/07860-9), T.N.O. (2020/04368-6) and B.S. (2016/50438-0) have been financially supported by Fundação de Amparo à Pesquisa do Estado de São Paulo. B.S. has also received a grant from Faculdade de Medicina da Universidade de São Paulo (2020.1.362.5.2).

The results of the study are presented clearly, honestly, and without fabrication, falsification, or inappropriate data manipulation.

